# Implementation of Point of Care HIV Viral load monitoring for people living with HIV in Low and Middle-income Countries: A systematic review on implementation research outcomes

**DOI:** 10.1101/2024.11.04.24316630

**Authors:** Perry Msoka, Iraseni Swai, Kennedy Ngowi, Ria Reis, Andreja Lekic, Blandina T. Mmbaga, Anita Hardon, Marion Sumari-de Boer

**Affiliations:** Kilimanjaro Clinical Research Institute, Moshi, Tanzania; Amsterdam Institute for Social Science Research, Amsterdam, the Netherlands; Amsterdam Institute for Global Health and Development, Amsterdam, the Netherlands; UMC Amsterdam, location AMC, Amsterdam, the Netherlands; The Children’s Institute, University of Cape Town, Cape Town, South Africa; Kilimanjaro Christian Medical University College, Moshi, Tanzania; Kilimanjaro Christian Medical Centre, Moshi, Tanzania; Knowledge, Technology and Innovation Chair group, Social Sciences Department, Wageningen University & Research, the Netherlands

**Keywords:** Point of Care HIV Viral Load monitoring, People living with HIV, systematic review, Implementation research outcomes, Low and middle-income countries

## Abstract

**Background:** Viral load monitoring has rapidly increased among people living with HIV(PLHIV) in low– and middle-income countries (LMICs), resulting in an increased laboratory workload. The use of innovative Point of Care (PoC) or near Point of Care (n)PoC HIV Viral Load (HIV VL) monitoring has enabled improved patient care, a reduction in laboratory workload, improved clinic retention and reduced turnaround time of results. However, implementation bottlenecks of such methods are uncertain, especially when PoC or (n)PoC is implemented in remote areas in low-volume clinics. The main aim of this study was to review implementation research outcomes of (n)PoC HIV VL monitoring for PLHIV in LMICs.

**Methodology:** We qualitatively synthesised peer-reviewed papers to explore implementation research outcomes (IROs) of (n)PoC HIV VL monitoring. We identified studies published between January 2013 and June 2024. We used the IROs described by Proctor et al., which are acceptability, adoption, appropriateness, cost, feasibility, fidelity, penetration and sustainability. We searched using the following Mesh terms: Point of care testing, HIV, viral load, acceptability, patient acceptance of health care, adoption, facilities and services utilisation, appropriateness, cost, feasibility, fidelity, penetration, coverage, sustainability and continuity of patient care through PubMed, Cochrane and Scopus. The PRISMA diagram in Figure 1 presents the selection process of included papers.

**Results:** Twenty-five studies reported implementation outcomes of PoC or (n)PoC HIV VL monitoring. Near PoC HIV VL monitoring using GeneXpert is considered acceptable to patients and healthcare providers. Point of care HIV VL monitoring using mPIMA was feasible as patients received the results the same day. From a health service provider’s perspective, PoC HIV VL monitoring was acceptable because it influenced patients to accept the illness and adhere to medication. Additionally, there was high testing coverage in routine PoC HIV VL monitoring centres. Fidelity was questionable in some settings due to (n)PoC HIV VL monitoring results not being delivered as intended. Additionally, we found in several studies that the (n)PoC costs are higher than standard of care test, USD 54.93 per test, at low testing volume clinics conducting 20VL tests per month compared to costs of USD 24.25 at high testing volume clinics conducting 100VL tests per month. However, costs are expected to be lower when (n)PoC HIV VL monitoring is scaled up and targeted for those at risk.

**Conclusion:** Implementation of PoC or (n)PoC testing for HIV viral load monitoring is acceptable and feasible and can reach a vast population. However, higher costs, limited fidelity, lower penetration and limited sustainability may hinder using (n)PoC testing in improving patient care and health outcomes. More knowledge and training should be implemented to overcome these challenges.

## Introduction

Globally, an estimated 39.9 million people were living with Human Immunodeficient Virus (HIV) in 2023, of whom 77% of adults and 57% of children accessed antiretroviral therapy (ART).(1) The Joint United Nations Programme on HIV and AIDS (UNAIDS) aims to reach 95-95-95 targets by 2030 that 95% of people living with HIV (PLHIV) know their HIV status, 95% of diagnosed PLHIV receive antiretroviral therapy (ART), and 95% of PLHIV receiving ART attain viral suppression below 1,000 copies/ml.(2) To reach these goals, the World Health Organization (WHO) emphasizes that all PLHIV receive ART and routine HIV viral load (VL) testing to monitor treatment response, adherence and virologic suppression.(3)

**Figure 1:**
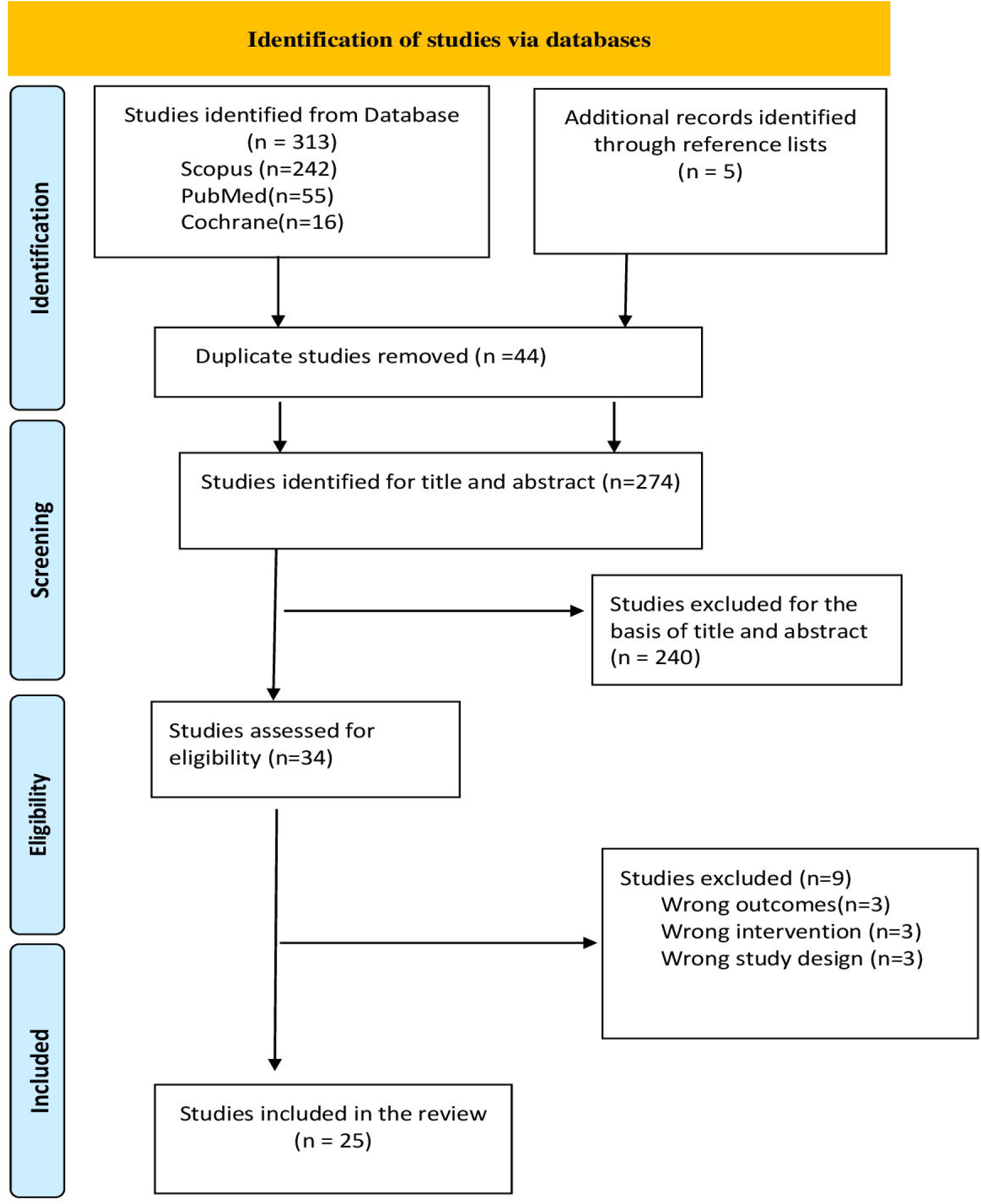
PRISMA diagram of studies screening and selection

However, monitoring the VL of PLHIV on ART in resource-limited settings is still challenging due to limited infrastructure, shortage of skilled staff, loss of samples, an inefficient system of providing results and patients being lost to follow-up.(4) To overcome those challenges, WHO has advocated for integrating Point of care HIV Viral load (PoC HIV VL) monitoring in HIV care services.(5,6)

Point of Care HIV VL monitoring entails testing at the point where healthcare is provided. The results can be given on the same day. Also, it can usually be performed by non-trained staff.(7) Near Point of Care HIV VL monitoring is testing in laboratories close to treatment facilities, while still improving the turnaround time from sampling to result.(8) The current PoC HIV VL assays that are commercially available or in late development include the PoC HIV assay of mPIMA by Abbott, and the (n)PoC HIV assays SAMBA by Diagnostics for the Real World and GeneXpert HIV 1 by Cepheid.(9)

Several studies have been conducted on the performance of PoC HIV VL monitoring. In Mozambique, mPIMA was found to have a high sensitivity of 95% and specificity of 96.5% to identify virological failure at the threshold of 1000 copies/ml when a standard conventional plasma test was used as the golden standard.(10) In addition, a study evaluating the performance of SAMBA in Malawi and Uganda found it to be accurate in differentiating patients who had a VL below or above 1000 copies/ml.(11)

Furthermore, a systematic review study from developed and developing countries found that PoC testing is preferable to centralized testing, with a sensitivity of 93.3%-100% and specificity of 99.5%-100% for Early Infant Diagnosis (EID), acute HIV infection diagnosis, or VL monitoring.(12)

Despite the good performance of PoC HIV VL monitoring in previous studies, several challenges exist. A systematic review of the performance and clinical utility of POC HIV viral load testing in resource-limited settings observed that there is a need for constant electricity for the machine to function and a temperature control room for sample processing and storage.(12)

Other reported challenges of PoC HIV VL monitoring are device operation by untrained staff, routine device maintenance, and the high costs of a PoC test. All these may hamper the implementation of PoC HIV VL monitoring.

Given the above-reported challenges, future studies should explore solutions to overcome possible implementation challenges. Proctor and colleagues (2011) designed a framework that describes eight distinct implementation outcomes to guide the successful implementation of innovations: acceptability, adoption, appropriateness, feasibility, fidelity, implementation costs, penetration and sustainability. Acceptability is the perception among stakeholders that a given treatment, service, practice, or innovation is agreeable, palatable, or satisfactory. Secondly, adoption is the intention, initial decision, or action to try or employ an innovation or evidence-based practice. Thirdly, appropriateness is the perceived fit of the innovation to address a particular issue or problem. Fourthly, the cost is the financial effect of an implementation effort. It may include charges for treatment delivery, implementation strategy and use of the service setting. Fifthly, feasibility is how a new treatment or innovation can be successfully used or carried out within a given agency or setting. Sixthly, fidelity is defined as the extent to which an intervention was implemented as prescribed in the original protocol or as the program developers intended. Seventhly, penetration is integrating practice within a service setting and its subsystems. Lastly, sustainability is how a newly implemented intervention is maintained or institutionalised within a service setting’s ongoing, stable operations. These implementation outcomes can give insights into challenges and assist in developing strategies to be investigated in future pragmatic trials.(13)

In this systematic review, we use the Proctor framework to explore implementation research outcomes of (n)PoC HIV VL monitoring for PLHIV in LMICs through a qualitative synthesis of peer-reviewed papers. The specific objectives were to review existing evidence on (1) the acceptability, (2) the adoption, (3) the appropriateness, (4) the cost, (5) the feasibility, (6) the fidelity, (7) the penetration and (8) the sustainability of (n)PoC HIV VL monitoring.

## Methodology

Using the reporting items in systematic review guideline(14) we qualitatively synthesised scientific papers to explore the implementation research outcomes (IROs) of PoC HIV VL monitoring for PLHIV. We used the Proctor framework of IROs and pre-defined topics to define the search terms.(13)

### Literature search

The initial broad relevant MESH terms were defined based on the terms ‘PoC testing’, ‘HIV’ and ‘Viral load’. We viewed the proper entry terminology for the IROs: Acceptability, Adoption, Appropriateness, Cost, Feasibility, Fidelity, Penetration and Sustainability. After defining these terms, we searched for the terms in June 2024 using a combination of the MESH and entry terms. A detailed search strategy is presented in the S1 Table.

Three databases were searched for publications published between January 2013 and June 2024:

(1) PubMed, (2) Cochrane and (3) Scopus. After this, we scanned the reference list of each obtained publication found through the above search for additional publications that could fit our search criteria.

### Study selection

Publications were included based on the following criteria:

- Quantitative, including trials or qualitative studies.
- Original papers.
- Focus on (n)PoC VL monitoring.
- A publication should at least describe one of the IROs as described by Proctor.

We used Covidence software https://support.covidence.org/help/how-can-i-cite-covidence to select and organise manuscripts and data extraction. The publications were identified and imported with a librarian (A) into the software for selecting eligible publications. After collecting publications from PubMed, Cochrane and Scopus, the titles and abstracts of identified studies were screened by two reviewers (PM, MS) for relevance. Any disagreements during the screening of titles and abstracts were resolved through discussion. In case of doubt that a publication met the criteria, it was included for full-text assessment to assess the eligibility. The full-text assessment of the quality and risk of bias of all included publications was performed by two reviewers (PM, MS). We used the Newcastle Ottawa scale (NOS) to assess the risk of bias. (15) The tool assessed the bias in three categories which are: selection, comparability, and outcome or exposure. A star system was used: a score of seven to nine stars was considered a low risk of bias, four to six stars were considered an “unclear risk of bias”, and three or fewer stars were considered a “high risk of bias”. The study was considered of low quality if we found it to have a high risk of bias.

All manuscripts included in the full-text assessment were judged based on the quality of the paper using twenty-seven criteria selected from the PRISMA guidelines for reporting systematic reviews.(16) The PRISMA checklist can be found in the S2 Table. The twenty selected criteria covered the following items:

- The abstract was comprehensive.
- The objective did clearly state the specific aims of the study.
- The rationale was based on a well-described literature review.
- The methodology clearly described qualitative or quantitative methods, research setting and participants, methods and data collection procedures.
- Ethical issues were assessed by judging informed consent procedures, voluntary participation, no harm, confidentiality and anonymity.
- The analysis and results had to be realistic to achieve the required study objective.

Exclusion criteria were:

- Studies with outcomes that were not relevant to our review.
- Studies describing interventions outside of our area of interest.
- Studies and reviews of papers published before 2013 in which PoC VL testing was not widely available or implemented.
- Protocol studies.

### Data extraction and management

One reviewer (PM) extracted the data. The following information was extracted from the included studies:

- Country where the study was performed
- Study design classified as clinical trials, cohort, qualitative or modelling studies
- Study funding sources
- Population description
- Study period, start and end date
- Inclusion and exclusion criteria
- Methods of recruitment of the participants
- Implementation of research outcomes (IROs) investigated

### Ethical Approval and Informed Consent

The review was registered on the PROSPERO registry for systematic reviews. (PROSPERO 2023 CRD42023394668). The review did not involve collecting new data from human subjects. Therefore, ethical approval and informed consent were not required.

## Results

Three hundred and thirteen studies were found through the initial search of PubMed, Scopus and Cochrane, and an additional five studies were found through scanning the reference lists. Forty-four duplicate studies were removed from the 318 studies. As a result, 274 studies were screened for eligibility based on title and abstract. From that, 240 irrelevant studies were excluded because the publications focused on something other than (n) PoC VL monitoring. Thirty-four studies were assessed on eligibility by reading the full text of the paper. We found that nine studies were not eligible: three had irrelevant outcomes for our review, three described interventions outside our area of interest, and three had wrong study designs, as detailed in the S3 Table. This led to 25 studies being included in the final review, as shown in Table 1. The PRISMA diagram showing detailed information on search results and the selection process is presented in S1 Figure.

**Table 1.**
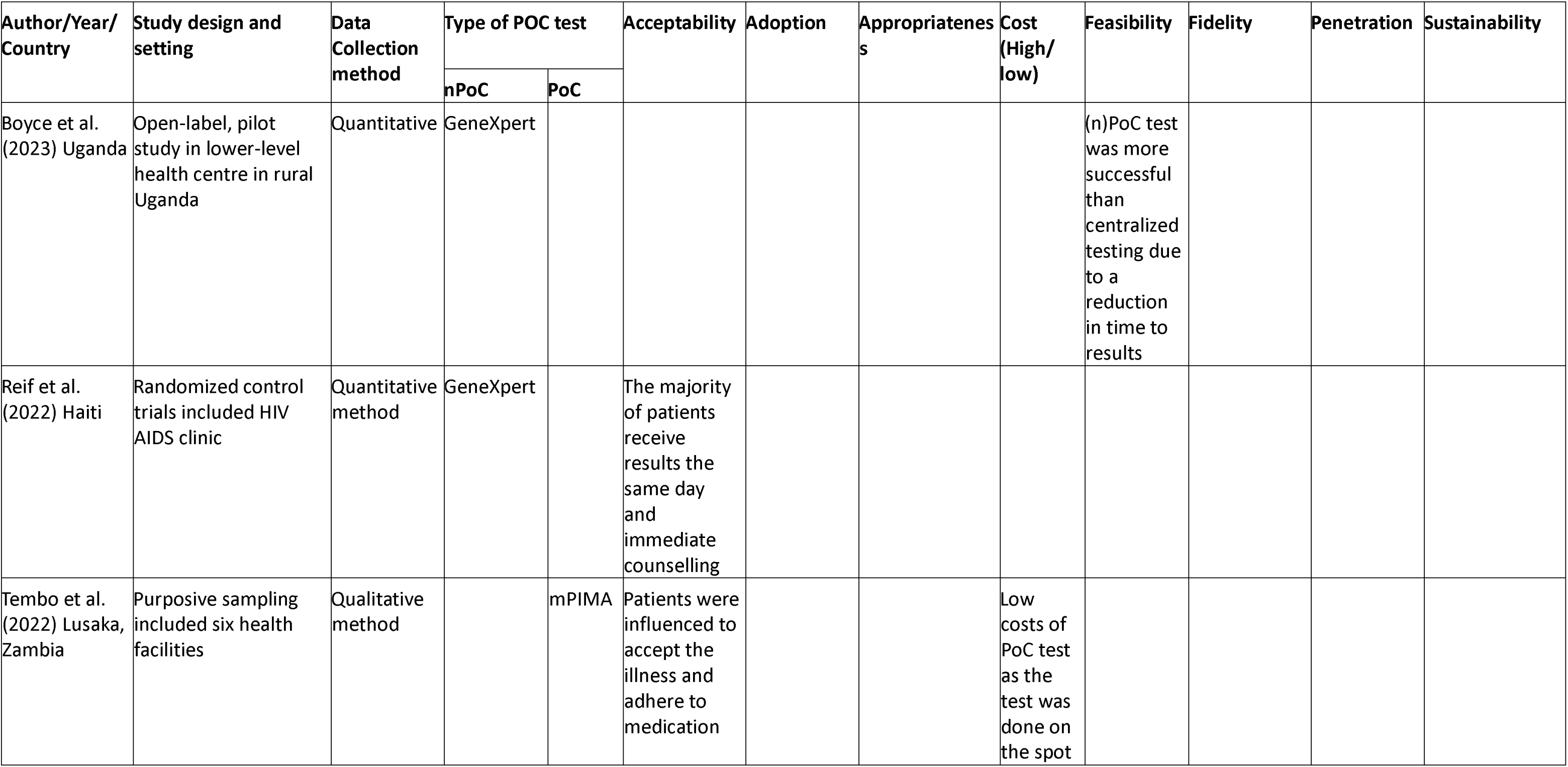
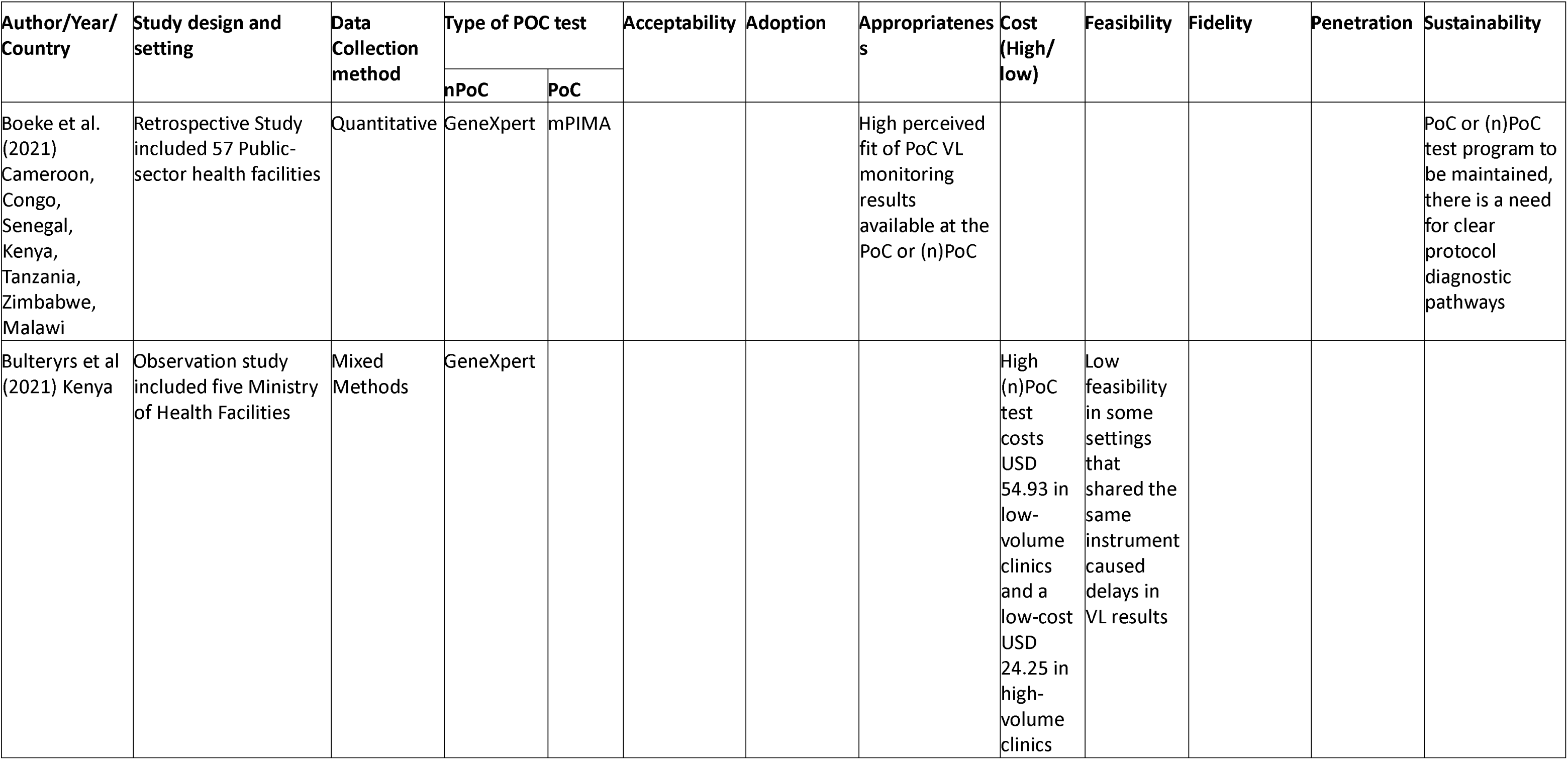
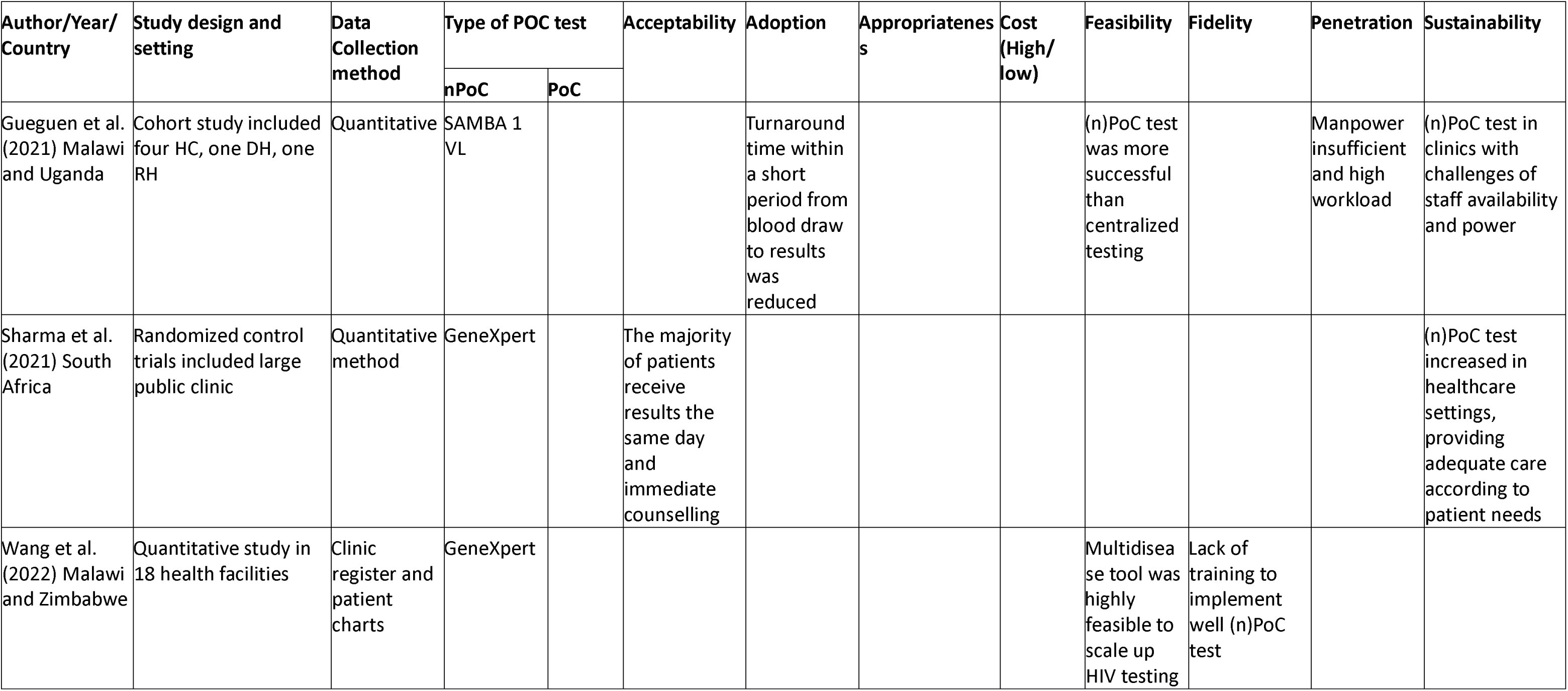
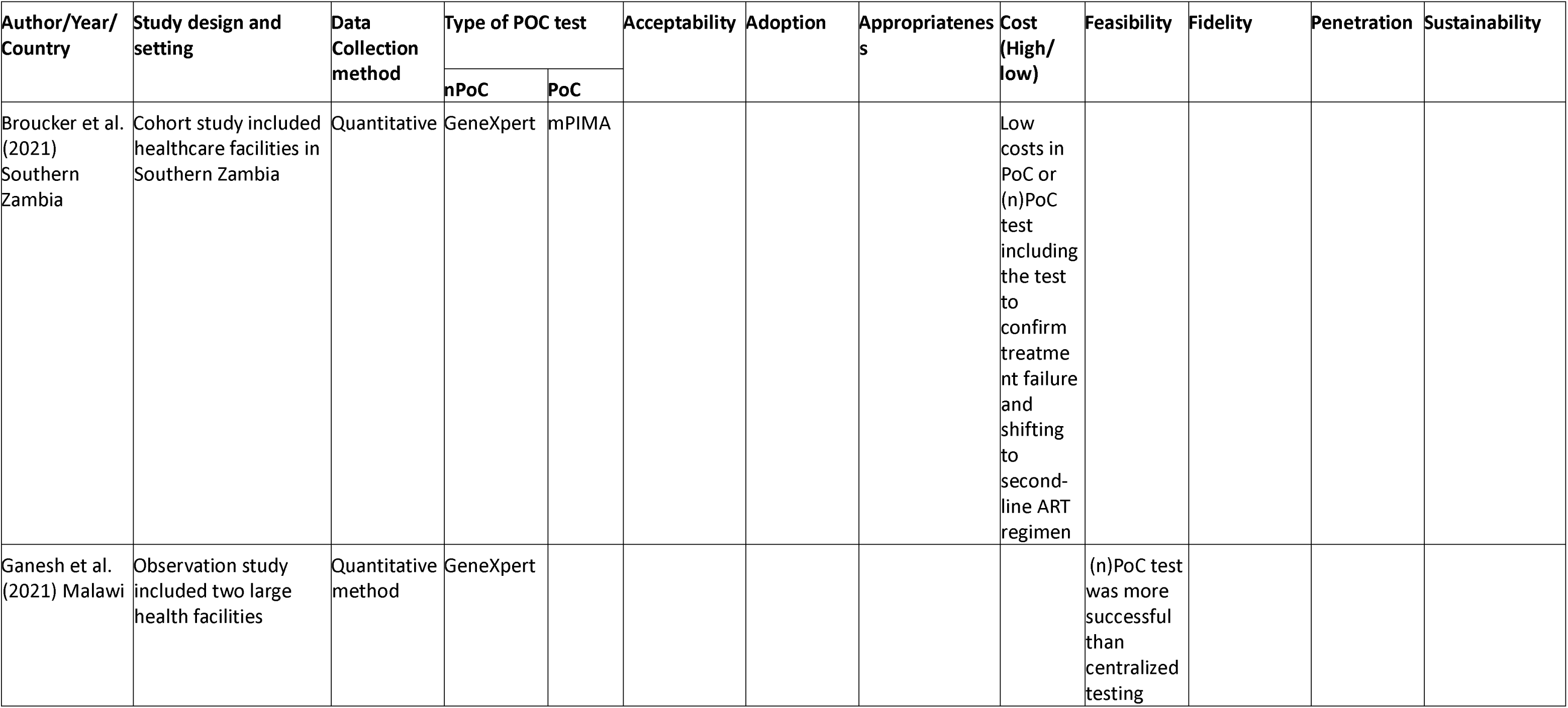
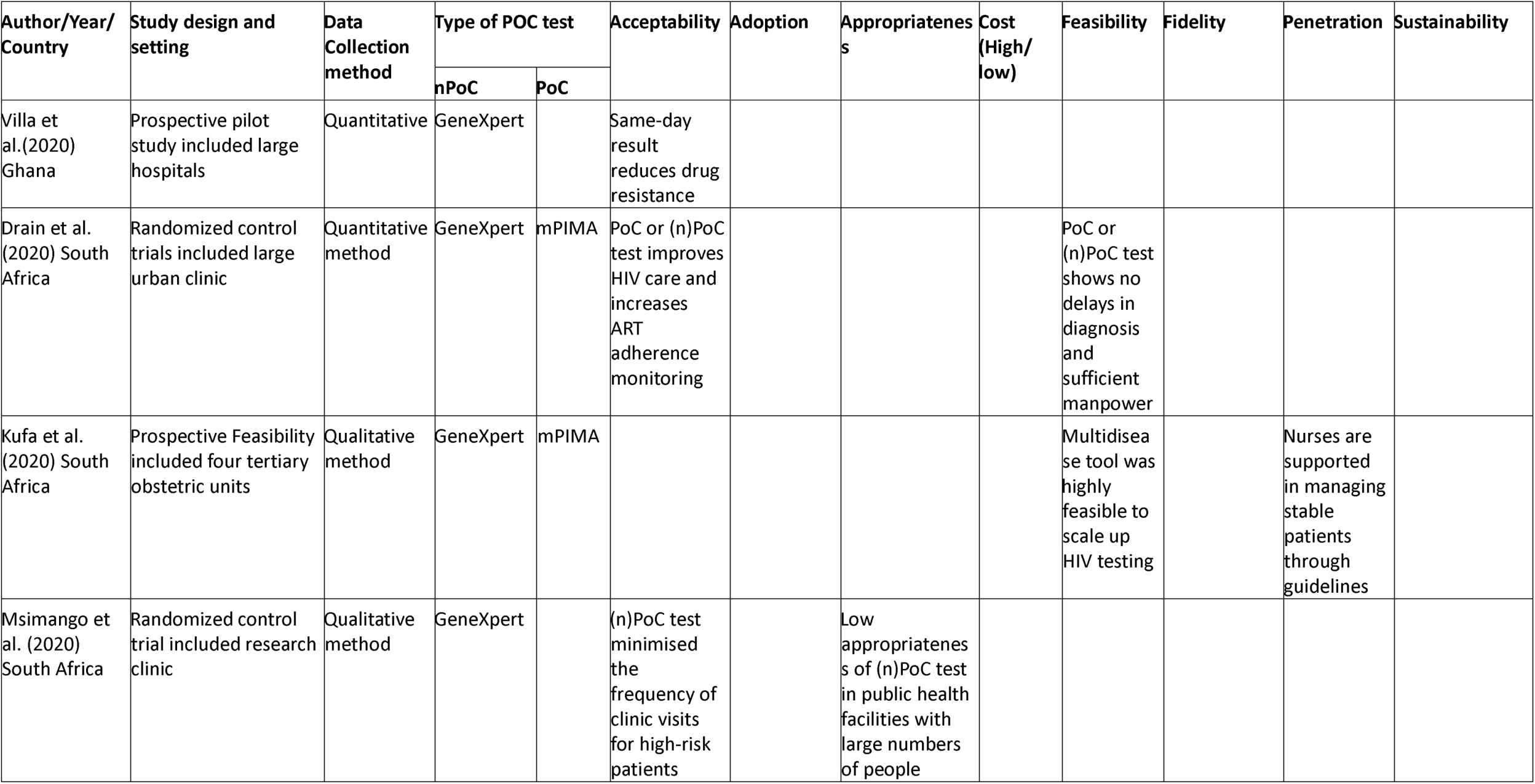
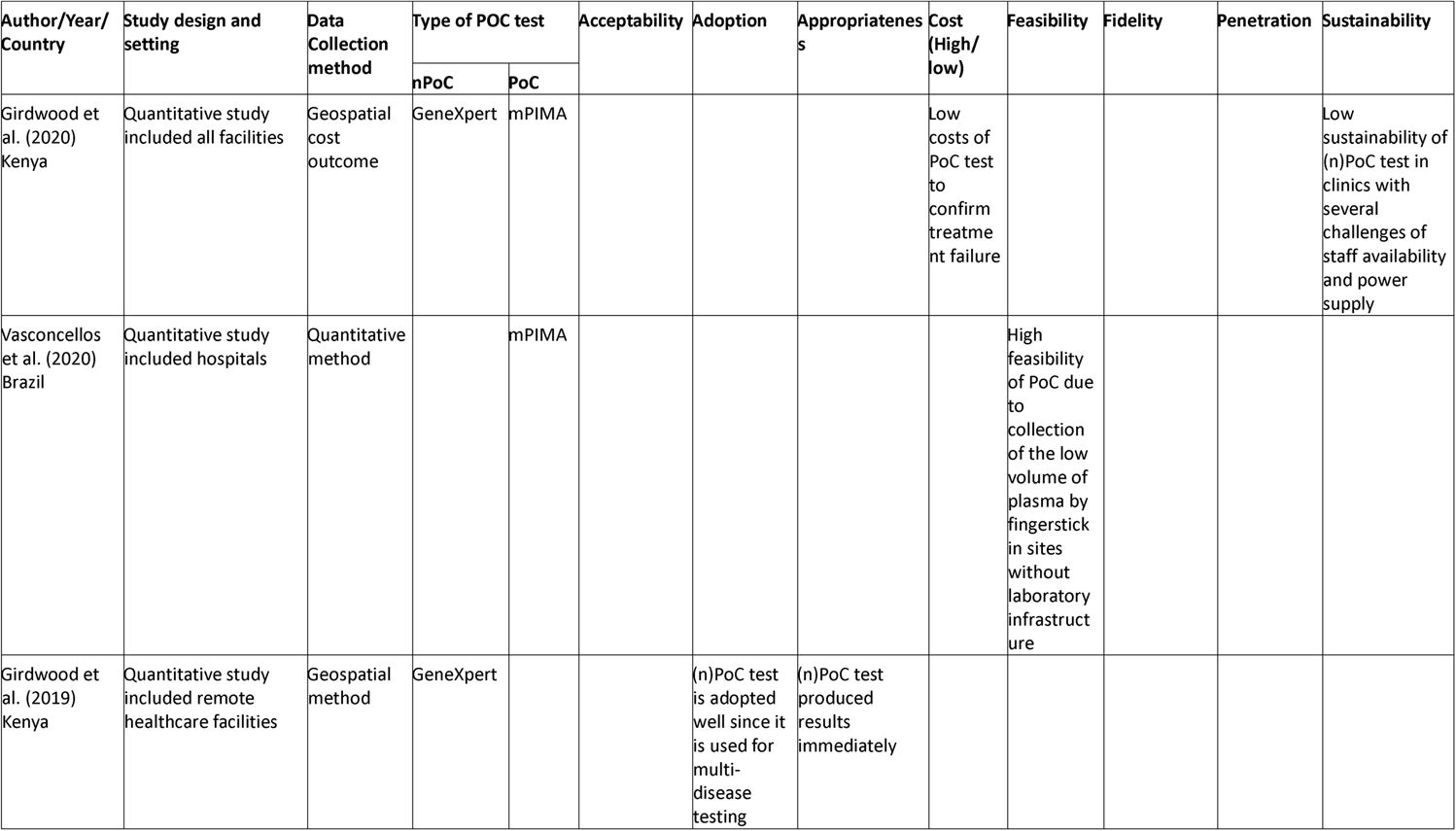
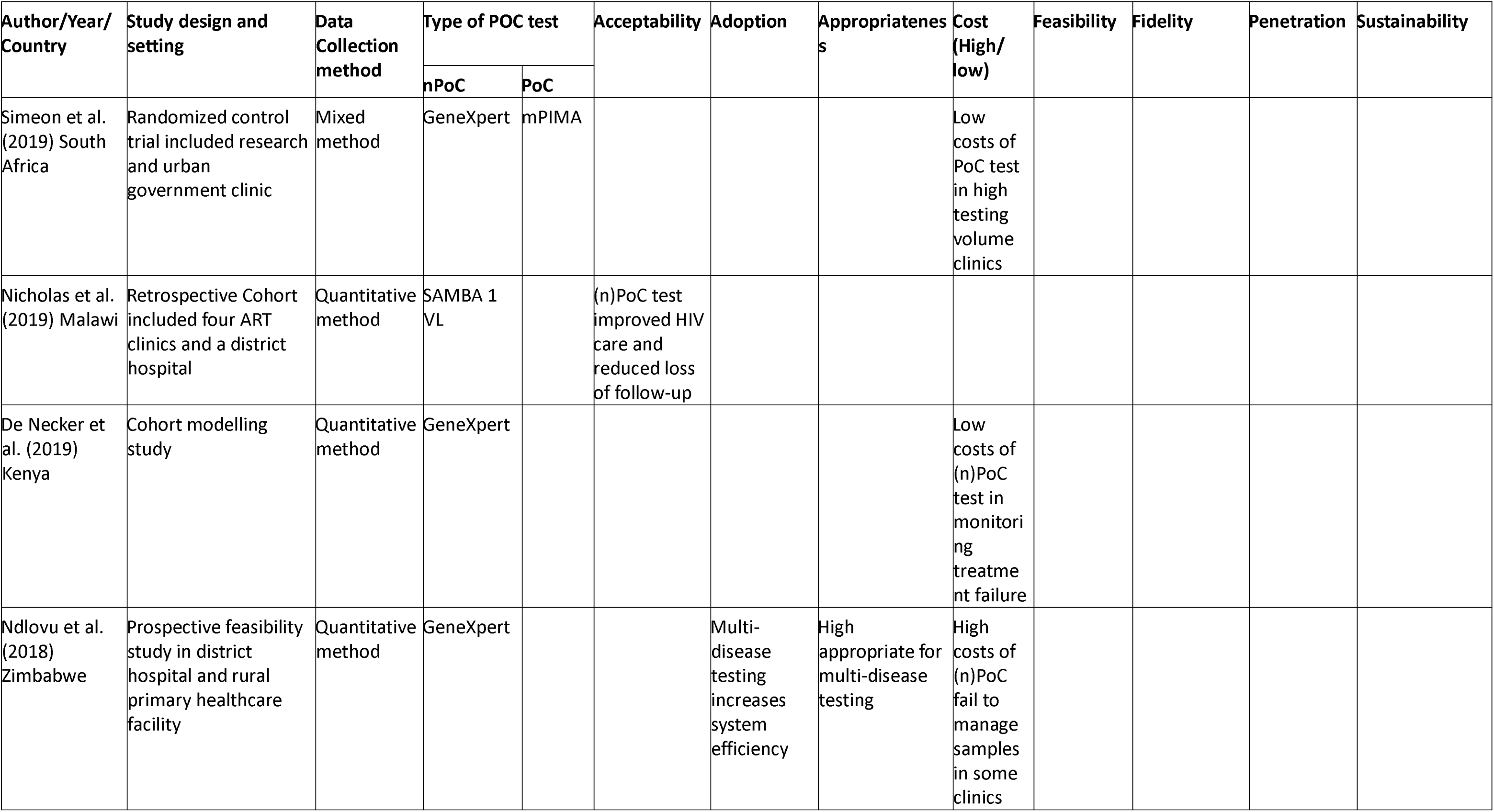
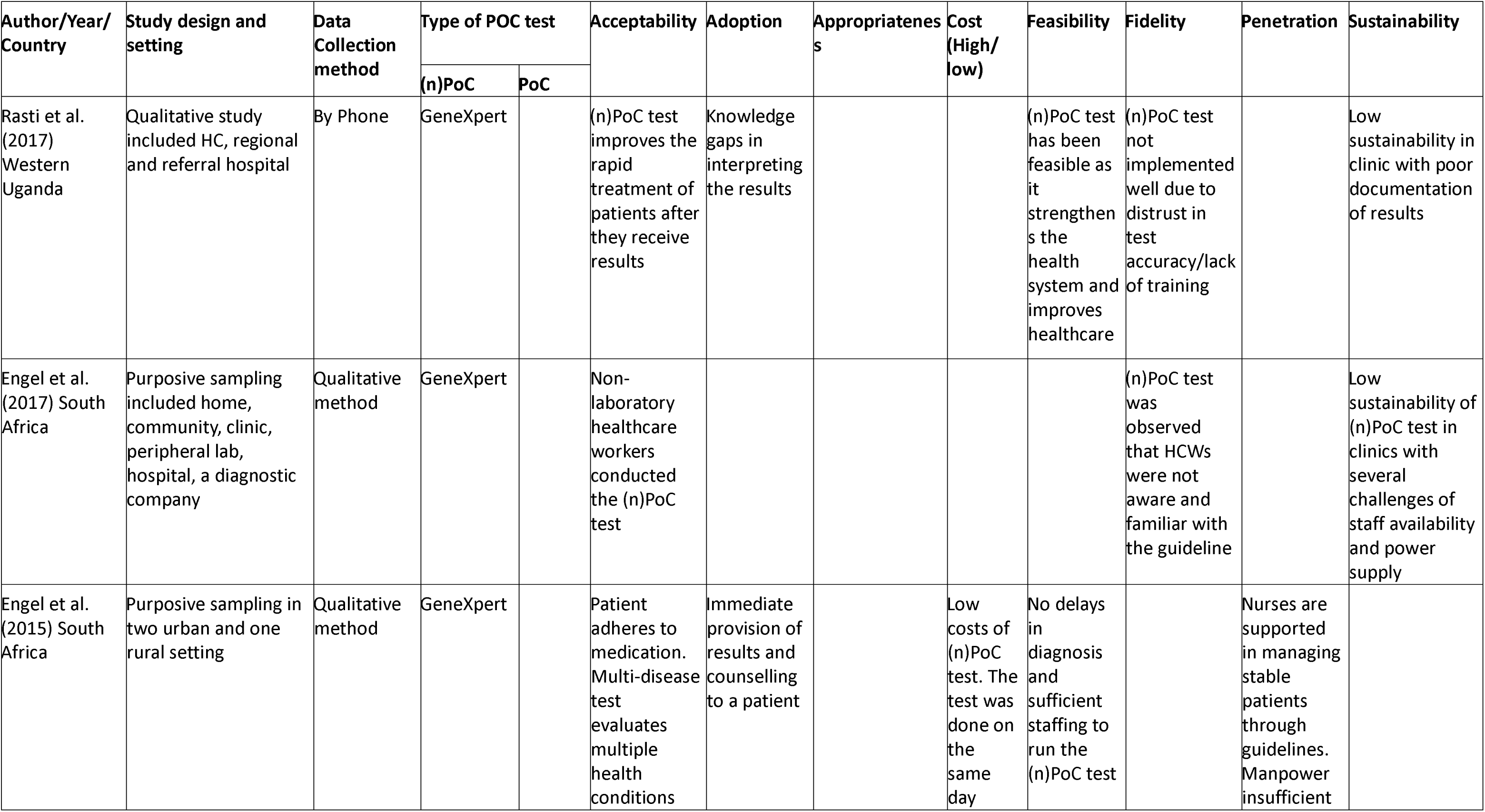
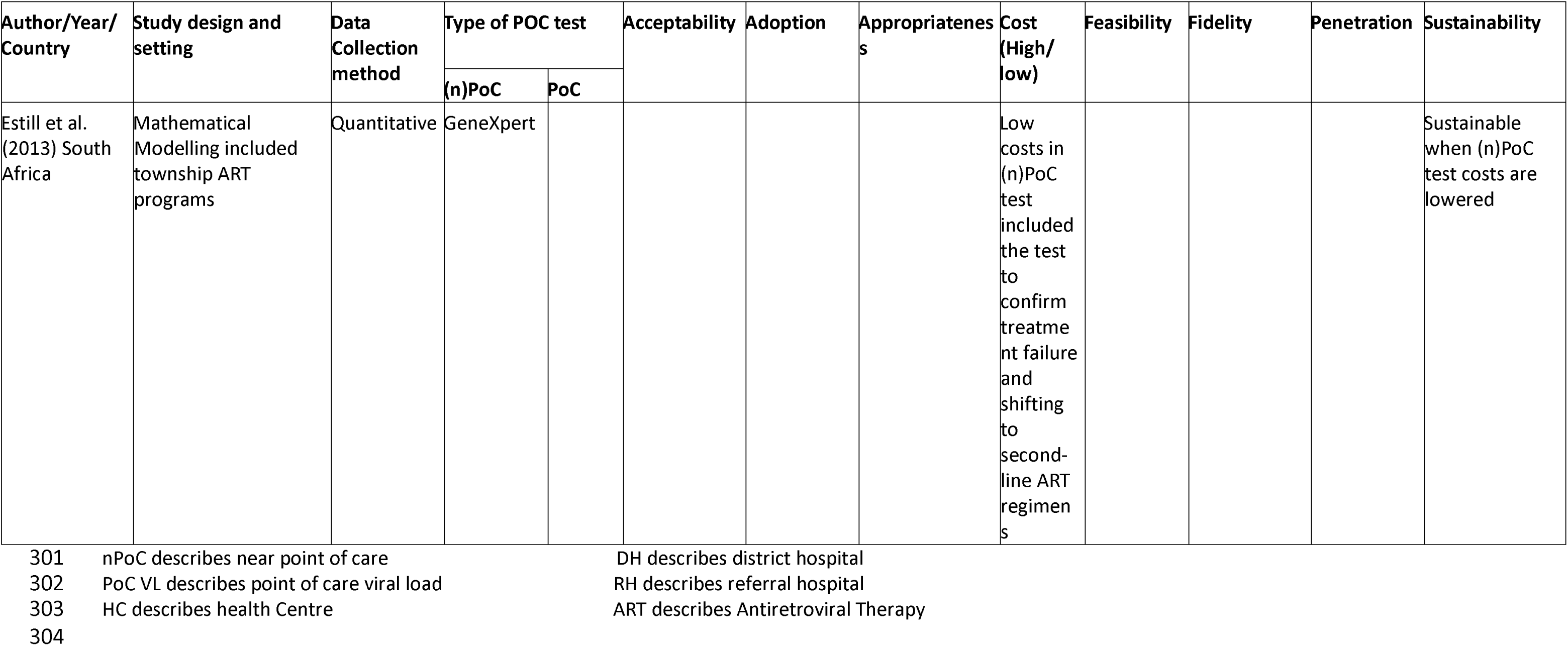
Characteristics of included studies.

| Author/Year/<br>Country | Study design and<br>setting | Data<br>Collection<br>method | Type of POC test |  | Acceptability | Adoption | Appropriatenes<br>s | Cost<br>(High/<br>low) | Feasibility | Fidelity | Penetration | Sustainability |
| --- | --- | --- | --- | --- | --- | --- | --- | --- | --- | --- | --- | --- |
|  |  |  | nPoC | PoC |  |  |  |  |  |  |  |  |
| Boyce et al.<br>(2023) Uganda | Open-label, pilot<br>study in lower-level<br>health centre in rural<br>Uganda | Quantitative | GeneXpert |  |  |  |  |  | (n)PoC test<br>was more<br>successful<br>than<br>centralized<br>testing due<br>to a<br>reduction<br>in time to<br>results |  |  |  |
| Reif et al.<br>(2022) Haiti | Randomized control<br>trials included HIV<br>AIDS clinic | Quantitative<br>method | GeneXpert |  | The majority<br>of patients<br>receive<br>results the<br>same day<br>and<br>immediate<br>counselling |  |  |  |  |  |  |  |
| Tembo et al.<br>(2022) Lusaka,<br>Zambia | Purposive sampling<br>included six health<br>facilities | Qualitative<br>method |  | mPIMA | Patients were<br>influenced to<br>accept the<br>illness and<br>adhere to<br>medication |  |  | Low<br>costs of<br>PoC test<br>as the<br>test was<br>done on<br>the spot |  |  |  |  |
|  |  |  | nPoC | PoC |  |  |  |  |  |  |  |  |
| Boeke et al.<br>(2021)<br>Cameroon,<br>Congo,<br>Senegal,<br>Kenya,<br>Tanzania,<br>Zimbabwe,<br>Malawi | Retrospective Study<br>included 57 Public-<br>sector health facilities | Quantitative | GeneXpert | mPIMA |  |  | High perceived<br>fit of PoC VL<br>monitoring<br>results<br>available at the<br>PoC or (n)PoC |  |  |  |  | PoC or (n)PoC<br>test program to<br>be maintained,<br>there is a need<br>for clear<br>protocol<br>diagnostic<br>pathways |
| Bulterys et al<br>(2021) Kenya | Observation study<br>included five Ministry<br>of Health Facilities | Mixed<br>Methods | GeneXpert |  |  |  |  | High<br>(n)PoC<br>test<br>costs<br>USD<br>54.93 in<br>low-volume<br>clinics<br>and a<br>low-cost<br>USD<br>24.25 in<br>high-<br>volume<br>clinics | Low<br>feasibility<br>in some<br>settings<br>that<br>shared the<br>same<br>instrument<br>caused<br>delays in<br>VL results |  |  |  |
|  |  |  | nPoC | PoC |  |  |  |  |  |  |  |  |
| Gueguen et al.<br>(2021) Malawi<br>and Uganda | Cohort study included<br>four HC, one DH, one<br>RH | Quantitative | SAMBA 1<br>VL |  |  | Turnaround<br>time within<br>a short<br>period from<br>blood draw<br>to results<br>was<br>reduced |  |  | (n)PoC test<br>was more<br>successful<br>than<br>centralized<br>testing |  | Manpower<br>insufficient<br>and high<br>workload | (n)PoC test in<br>clinics with<br>challenges of<br>staff availability<br>and power |
| Sharma et al.<br>(2021) South<br>Africa | Randomized control<br>trials included large<br>public clinic | Quantitative<br>method | GeneXpert |  | The majority<br>of patients<br>receive<br>results the<br>same day<br>and<br>immediate<br>counselling |  |  |  |  |  |  | (n)PoC test<br>increased in<br>healthcare<br>settings,<br>providing<br>adequate care<br>according to<br>patient needs |
| Wang et al.<br>(2022) Malawi<br>and Zimbabwe | Quantitative study in<br>18 health facilities | Clinic<br>register and<br>patient<br>charts | GeneXpert |  |  |  |  |  | Multidisea<br>se tool was<br>highly<br>feasible to<br>scale up<br>HIV testing | Lack of<br>training to<br>implement<br>well (n)PoC<br>test |  |  |
|  |  |  | nPoC | PoC |  |  |  |  |  |  |  |  |
| Broucker et al.<br>(2021)<br>Southern<br>Zambia | Cohort study included<br>healthcare facilities in<br>Southern Zambia | Quantitative | GeneXpert | mPIMA |  |  |  | Low<br>costs in<br>PoC or<br>(n)PoC<br>test<br>including<br>the test<br>to<br>confirm<br>treatme<br>nt failure<br>and<br>shifting<br>to<br>second-line ART<br>regimen |  |  |  |  |
| Ganesh et al.<br>(2021) Malawi | Observation study<br>included two large<br>health facilities | Quantitative<br>method | GeneXpert |  |  |  |  |  | (n)PoC test<br>was more<br>successful<br>than<br>centralized<br>testing |  |  |  |
|  |  |  | nPoC | PoC |  |  |  |  |  |  |  |  |
| Villa et al.(2020)<br>Ghana | Prospective pilot study included large hospitals | Quantitative | GeneXpert |  | Same-day result reduces drug resistance |  |  |  |  |  |  |  |
| Drain et al. (2020) South Africa | Randomized control trials included large urban clinic | Quantitative method | GeneXpert | mPIMA | PoC or (n)PoC test improves HIV care and increases ART adherence monitoring |  |  |  | PoC or (n)PoC test shows no delays in diagnosis and sufficient manpower |  |  |  |
| Kufa et al. (2020) South Africa | Prospective Feasibility included four tertiary obstetric units | Qualitative method | GeneXpert | mPIMA |  |  |  |  | Multidisease tool was highly feasible to scale up HIV testing |  | Nurses are supported in managing stable patients through guidelines |  |
| Msimango et al. (2020) South Africa | Randomized control trial included research clinic | Qualitative method | GeneXpert |  | (n)PoC test minimised the frequency of clinic visits for high-risk patients |  | Low appropriatenes of (n)PoC test in public health facilities with large numbers of people |  |  |  |  |  |

| Author/Year/<br>Country | Study design and<br>setting | Data<br>Collection<br>method | Type of POC test |  | Acceptability | Adoption | Appropriateness | Cost<br>(High/<br>low) | Feasibility | Fidelity | Penetration | Sustainability |
| --- | --- | --- | --- | --- | --- | --- | --- | --- | --- | --- | --- | --- |
|  |  |  | nPoC | PoC |  |  |  |  |  |  |  |  |
| Girdwood et al. (2020)<br>Kenya | Quantitative study included all facilities | Geospatial cost outcome | GeneXpert | mPIMA |  |  |  | Low costs of PoC test to confirm treatment failure |  |  |  | Low sustainability of (n)PoC test in clinics with several challenges of staff availability and power supply |
| Vasconcellos et al. (2020)<br>Brazil | Quantitative study included hospitals | Quantitative method |  | mPIMA |  |  |  |  | High feasibility of PoC due to collection of the low volume of plasma by fingerstick in sites without laboratory infrastructure |  |  |  |
| Girdwood et al. (2019)<br>Kenya | Quantitative study included remote healthcare facilities | Geospatial method | GeneXpert |  |  | (n)PoC test is adopted well since it is used for multi-disease testing | (n)PoC test produced results immediately |  |  |  |  |  |
|  |  |  | nPoC | PoC |  |  |  |  |  |  |  |  |
| Simeon et al.<br>(2019) South<br>Africa | Randomized control<br>trial included research<br>and urban<br>government clinic | Mixed<br>method | GeneXpert | mPIMA |  |  |  | Low<br>costs of<br>PoC test<br>in high<br>testing<br>volume<br>clinics |  |  |  |  |
| Nicholas et al.<br>(2019) Malawi | Retrospective Cohort<br>included four ART<br>clinics and a district<br>hospital | Quantitative<br>method | SAMBA 1<br>VL |  | (n)PoC test<br>improved HIV<br>care and<br>reduced loss<br>of follow-up |  |  |  |  |  |  |  |
| De Necker et<br>al. (2019)<br>Kenya | Cohort modelling<br>study | Quantitative<br>method | GeneXpert |  |  |  |  | Low<br>costs of<br>(n)PoC<br>test in<br>monitori<br>ng<br>treatme<br>nt failure |  |  |  |  |
| Ndlovu et al.<br>(2018)<br>Zimbabwe | Prospective feasibility<br>study in district<br>hospital and rural<br>primary healthcare<br>facility | Quantitative<br>method | GeneXpert |  |  | Multi-disease<br>testing<br>increases<br>system<br>efficiency | High<br>appropriate for<br>multi-disease<br>testing | High<br>costs of<br>(n)PoC<br>fail to<br>manage<br>samples<br>in some<br>clinics |  |  |  |  |

| Author/Year/<br>Country | Study design and<br>setting | Data<br>Collection<br>method | Type of POC test |  | Acceptability | Adoption | Appropriateness | Cost<br>(High/<br>low) | Feasibility | Fidelity | Penetration | Sustainability |
| --- | --- | --- | --- | --- | --- | --- | --- | --- | --- | --- | --- | --- |
|  |  |  | (n)PoC | PoC |  |  |  |  |  |  |  |  |
| Rasti et al.<br>(2017)<br>Western<br>Uganda | Qualitative study<br>included HC, regional<br>and referral hospital | By Phone | GeneXpert |  | (n)PoC test<br>improves the<br>rapid<br>treatment of<br>patients after<br>they receive<br>results | Knowledge<br>gaps in<br>interpreting<br>the results |  |  | (n)PoC test<br>has been<br>feasible as<br>it<br>strengthens<br>the<br>health<br>system and<br>improves<br>healthcare | (n)PoC test<br>not<br>implemented<br>well due to<br>distrust in<br>test<br>accuracy/lack<br>of training |  | Low<br>sustainability in<br>clinic with poor<br>documentation<br>of results |
| Engel et al.<br>(2017) South<br>Africa | Purposive sampling<br>included home,<br>community, clinic,<br>peripheral lab,<br>hospital, a diagnostic<br>company | Qualitative<br>method | GeneXpert |  | Non-laboratory<br>healthcare<br>workers<br>conducted<br>the (n)PoC<br>test |  |  |  |  | (n)PoC test<br>was<br>observed<br>that HCWs<br>were not<br>aware and<br>familiar with<br>the guideline |  | Low<br>sustainability of<br>(n)PoC test in<br>clinics with<br>several<br>challenges of<br>staff availability<br>and power<br>supply |
| Engel et al.<br>(2015) South<br>Africa | Purposive sampling in<br>two urban and one<br>rural setting | Qualitative<br>method | GeneXpert |  | Patient<br>adheres to<br>medication.<br>Multi-disease<br>test<br>evaluates<br>multiple<br>health<br>conditions | Immediate<br>provision of<br>results and<br>counselling<br>to a patient |  | Low<br>costs of<br>(n)PoC<br>test. The<br>test was<br>done on<br>the<br>same<br>day | No delays<br>in<br>diagnosis<br>and<br>sufficient<br>staffing to<br>run the<br>(n)PoC test |  | Nurses are<br>supported<br>in managing<br>stable<br>patients<br>through<br>guidelines.<br>Manpower<br>insufficient |  |
|  |  |  | (n)PoC | PoC |  |  |  |  |  |  |  |  |
| Estill et al.<br>(2013) South<br>Africa | Mathematical<br>Modelling included<br>township ART<br>programs | Quantitative | GeneXpert |  |  |  |  | Low<br>costs in<br>(n)PoC<br>test<br>included<br>the test<br>to<br>confirm<br>treatme<br>nt failure<br>and<br>shifting<br>to<br>second-line ART<br>regimen<br>s |  |  |  | Sustainable<br>when (n)PoC<br>test costs are<br>lowered |

nPoC describes near point of care
PoC VL describes point of care viral load
HC describes health Centre
DH describes district hospital
RH describes referral hospital
ART describes Antiretroviral Therapy

### Study characteristics

Twenty-five papers were included in this systematic review.(17–41)Among the studies, seventeen were quantitative studies,(18,19,21,22,24–28,30–32,34,36–38,41) six were qualitative studies,(20,29,33,35,39,40) and two were mixed methods studies.(17,23)

### Overall results

We assessed the risk of bias with the Newcastle Ottawa Scale.(15) Fourteen studies had a low risk of bias (scored 7-9).(17,18,21–25,28–32,35,38) while ten studies had an unclear risk of bias (scored 4-6).(19,20,26,27,33,34,36,37,39,40) Only one study had a high risk of bias and scored 3; this was the prospective feasibility study conducted in Zimbabwe that highlighted potential bottlenecks.(41).The Newcastle Ottawa Scale completed in detail is available in the S4 Table.

Each study described at least one of the eight IROs defined by Proctor et al.(13)

### Acceptability

Ten studies described the acceptability of (n)PoC VL monitoring.(18–21,25,29,33,35,38,40) A study in Malawi, Uganda and South Africa (SA) on SAMBA I HIV, reported high satisfaction with (n)PoC VL monitoring, reduced turnaround times from blood draw to results and at the same time minimizing clinic visits for high-risk patients.(38,40) Further, in the STREAM study in SA, a retrospective cohort study on SAMBA 1 VL in Malawi, and a qualitative study in Southwestern Uganda, PoC or (n)PoC VL monitoring was highly acceptable to patients as it improves HIV care and ART adherence monitoring.(18–20)

In addition, studies conducted in Ghana and Haiti using GeneXpert showed that (n)PoC VL monitoring was highly acceptable to patients with the added benefit of results on the same day, reduced drug resistance and immediate counselling.(21,25) Likewise, a qualitative study in SA and Zambia showed that (n)PoC or PoC VL monitoring was highly acceptable by health providers as it helps patients to accept their illness and adhere to medication.(29,33) However, low acceptability of (n)PoC VL monitoring was observed in the SA studies as nonlaboratory healthcare workers conducted (n)PoC tests.(29,35)

### Adoption

Four studies described the adoption of (n)PoC VL monitoring.(20,29,30,41) Medical professionals and patients observed a high uptake of (n)PoC VL monitoring in studies conducted in SA, Kenya and Zimbabwe for its immediate provision of results and counselling to patients, including multi-disease testing, which increases system efficiency.(29,30,41) However, in a qualitative study conducted in Southwestern Uganda, health staff observed low adoption of (n)PoC VL monitoring, coupled with limited knowledge regarding interpreting the results.(20)

### Appropriateness

Appropriateness was touched upon in four studies.(30,32,40,41) A retrospective study in seven Sub-Saharan countries and the STREAM study in SA showed high appropriateness of (n)PoC VL monitoring as results were available at the near point of care.(32,40)

In addition, a quantitative study in Kenya and the prospective feasibility study conducted in Zimbabwe revealed that (n)PoC testing with GeneXpert was highly appropriate for multi-disease testing as it provides immediate results(30,41) On the contrary, the second qualitative study done in SA observed a low appropriateness of (n)PoC VL monitoring in public health facilities with large numbers of people.(40)

### Costs

Costs were described in nine studies.(17,23,27–29,31,33,36,41) An observational study of (n)PoC VL monitoring conducted in Kenya revealed high costs in low-testing volume clinics.(17) Researchers compared the (n)PoC costs with centralised testing, which were 54.93 USD per test and 25.65 USD per test respectively. The costs include personal time for a blood draw, clinical supplies, lab processing, training, lab equipment, electricity, rental space and cartridges. Furthermore, the feasibility study in Zimbabwe showed that (n)PoC VL monitoring had high costs due to failure to manage samples in some clinics.(41)

A cohort study in SA and Zambia and quantitative studies in Kenya observed low costs of PoC VL monitoring that included both the confirmation of treatment failure and the transition to second-line ART regimens.(27,28,31,36) Also, qualitative studies in SA and Zambia showed low costs of PoC tests, as it was performed on the same day and on the spot.(29,33) In addition, a mixed method study in SA showed low costs of (n)PoC VL monitoring in high testing volume clinics.(23)

### Feasibility

Feasibility was described in ten studies.(17,18,20,24,26,29,34,37–39) The SAMBA and feasibility studies in Malawi and Uganda using GeneXpert found that (n)PoC VL monitoring was more successful due to a reduction in time to results than centralised testing.(34,37,38)

Similarly, a prospective study on the implementation of PoC maternal VL and Early infant diagnosis conducted in SA and the feasibility study in Malawi and Zimbabwe showed that GeneXpert used as a multi-disease testing tool, was highly feasible to scale up HIV testing.(26,39) Furthermore, the STREAM and the qualitative studies in SA and Southwestern Uganda found high feasibility of (n)PoC VL monitoring, enabling rapid diagnosis, which, when coupled with sufficient staffing, contributes to the improvement of healthcare.(18,20,29)

Similarly, the quantitative study on developing and validating a simple and rapid way to generate a low volume of plasma used in PoC VL technology observed high feasibility in sites without laboratory Infrastructure.(24) However, low feasibility was observed in the Kenyan observational study in some settings that shared the same instrument, causing delays in VL results.(17)

### Fidelity

Fidelity was described in three studies. (20,26,35) The qualitative study in Southwestern Uganda and the feasibility study in Malawi and Zimbabwe revealed that (n)PoC VL monitoring was not implemented accurately. This was due to clients’ distrust in the accuracy of the tests and insufficient training for HCWs.(20,26) Also, the qualitative study in SA showed a low fidelity of (n)PoC VL monitoring as HCWs were not aware or familiar with the guidelines.(35)

### Penetration

Three studies reported penetration.(29,38,39) The qualitative, prospective studies in SA reported high penetration of PoC or (n)PoC services when nurses are supported in managing stable patients through guidelines.(29,39) On the contrary, a qualitative study in SA and the SAMBA study in Malawi and Uganda observed low penetration of the (n)PoC VL monitoring programme due to insufficient manpower and a high workload.(29,38)

### Sustainability

Sustainability was described in six studies.(20,22,32,35,36,38) A retrospective study in Sub-Saharan African countries, STREAM and cost-effective studies in SA observed high sustainability of PoC or (n)PoC VL monitoring, by having clear diagnostic pathways in healthcare settings, ensuring increased utilisation and adequate patient care, especially when lowering PoC test costs.(22,32,36)

On the other hand, the qualitative studies in Southwestern Uganda and SA and SAMBA studies in Malawi and Uganda observed low sustainability of (n)PoC VL monitoring in clinics, which is attributed to poor documentation of results, staff availability challenges and power supply.(20,35,38)

## Discussion

This review has accumulated valuable insights into how PoC VL monitoring was implemented in various settings in LMICs and how specific implementation bottlenecks may limit proper implementation. Out of the twenty-five studies, fifteen studies specifically used GeneXpert for (n)PoC VL monitoring, six studies employed both GeneXpert and mPIMA, two studies used SAMBA 1 VL and only two studies focused on mPIMA PoC VL monitoring.

The implementation outcomes of (n)PoC VL monitoring have been widely positive in most of the studies, with reported high satisfaction among patients and HCWs. The benefits reported include reduced turnaround time, minimised clinic visits for high-risk patients, improved HIV care and increased adherence monitoring to antiretroviral therapy (ART), which significantly contribute to the acceptance and uptake of (n)PoC VL monitoring.

The short turnaround time of (n)PoC VL monitoring, providing same-day results was a key factor in its high acceptability. This led to early detection of treatment failure, minimised clinic visits, improved patient outcomes and minimised drug resistance. This was observed in a study conducted by Msimango et al., that (n)PoC VL monitoring was preferred and satisfactory over central laboratory testing as it reduced the frequency of clinic visits. (41)

This review shows that (n)PoC VL monitoring has improved the quality of HIV care from the patient’s perspective. Additionally, a study by Nicholas et al., with SAMBA 1 VL test, showed that the same-day VL load test results improved HIV care and reduced follow-up loss.(19) Moreover, PoC or (n)PoC VL monitoring has shown to lead to increased adherence monitoring to ART. This was evident from a study in South Africa by Drain et al., in which patients highlighted the advantages of MPIMA PoC or GeneXpert (n)PoC VL monitoring and that it enhances the monitoring of adherence to Antiretroviral Treatment (ART).(18)

However, the adoption of PoC VL monitoring in various studies has been hindered by several implementation bottlenecks. Challenges include issues related to non-laboratory HCWs not knowing how to interpret the results well, making it difficult to integrate (n)PoC VL monitoring into healthcare practices. This was also noted in a study by Wang et al., where HCWs lacked training to implement (n)PoC VL monitoring tests effectively.(26)

Challenges also arose in terms of appropriateness in public health facilities, with large patient volumes leading to delays in clinic flow. According to a study by Msimango et al., challenges arose for the client, as (n)PoC tests may cause delays in clinic flow due to a high number of people in the facility.(40)

Costs emerged as a barrier, particularly in low-volume clinics due to the high costs of (n)PoC VL monitoring. The study by Simeon et al., noted that (n)PoC VL monitoring reduced its costs in low-volume clinics when used for other disease testing as well. (23)

However, the complexity of multi-disease testing was identified as a challenge too in the review studies. This was also reported by Bulterys et al., indicating that clinics with multi-disease testing for (n)PoC VL monitoring using GeneXpert are accompanied by several challenges, including multiple patient queues causing delays in results and extended clinic visits.(17) Implementation of (n)PoC VL monitoring challenges including test accuracy, were identified. Research by Rasti et al., found that patients were concerned about the diagnostic accuracy of (n)PoC VL monitoring.(20)

Insufficient manpower and high workloads were recognised as obstacles. Engel et al. reported that nurses perceived (n)PoC implementation as an extra workload, leading to patients’ delays in waiting for the results.(29)

To overcome these implementation bottlenecks, clear diagnostic pathways and proper documentation of results were identified as essential measures to ensure the successful implementation of PoC or (n)PoC VL monitoring.

Also, studies showed that (n)PoC VL monitoring was acceptable and well adopted by patients and HCWs due to multi-disease testing and delivery of same-day results.(21,30) On the other hand, PoC VL monitoring was shown to be cost-effective, appropriate and feasible when conducted at the point of care.(24,32,33)

This review has several limitations. The author considered only peer-reviewed published articles, did not include unpublished reviews and the period for the included studies was restricted. Studies published between 2013-2024 and limited databases were covered. The lack of data from geographic regions outside of low– and middle-income countries raises a concern about bias and the generalizability of the findings.

The strength of this review is that we used the Proctor framework, which gives a comprehensive overview of implementation research outcomes. Gaps could suggest further research that can provide better insights into using PoC or (n)PoC VL monitoring in healthcare settings, human resources, continuous staff training and sustainable supply chain of PoC or (n)PoC facilities.

### Recommendations

Point of Care VL monitoring provides rapid results followed by adherence counselling compared to nPoC VL monitoring. Rapid results provided by PoC VL monitoring improved its acceptability, leading to early detection of treatment failure, minimized clinic visits and improved patient outcomes. To address the implementation challenges of (n)PoC VL monitoring, prioritisation of clear diagnostic pathways and proper documentation are needed, as identified in this review. Further research should address gaps in healthcare settings, human resources, staff training and the sustainable supply chain for facilities that use (n)PoC tests.

## Conclusion

Implementing PoC or (n)PoC testing for HIV VL monitoring was acceptable, feasible and can reach a broad population. However, high costs, limited fidelity, and lack of penetration and sustainability may hinder the use of the (n)PoC test in improving patient care and health outcomes. Our findings indicate different implementation bottlenecks, such as non-laboratory HCWs not knowing how to interpret results and clinic delays due to high patient volumes. Additional challenges include increased costs due to low attendance, complex multi-disease testing and test accuracy concerns. Insufficient manpower and high workloads also affect implementation.

Point of care knowledge and training are needed to address problems related to implementation outcomes. This enables researchers, public health communities and policymakers to collaborate effectively in developing and testing evidence-based interventions.

## Data Availability

All relevant data are within the manuscript and its supporting information files

## Acknowledgements

The authors thank the members of the EAPOC-VL study. The study is a collaboration between Uganda National Health Research Organisation/Uganda Virus Research Institute (UNHRO/UVRI, Uganda), Karolinska Institute (Sweden), The Good Samaritan Foundation Kilimanjaro Christian Medical Centre (GSF KCMC, Tanzania), Stichting Amsterdam Institute for Global Health and Development (AIGHD, the Netherlands), London School of Hygiene and Tropical Medicine (MRC/UVRI & LSHTM Uganda Research Unit, Uganda), National Institute for Medical Research (NIMR, Tanzania), Kenya Medical Research Institute (KEMRI), Center for Global Health Research (CGHR), and the University of Rwanda (UR, Rwanda).

## Supporting information

S1 Figure PRISMA Diagram

S1 Table Search Strategy

S2 Table PRISMA checklists

S3 Table Reasons for exclusions

S4 Table Newcastle Ottawa Scale

## Authors’ contributions

PM performed the literature search, screening, data extraction and analysis and constructed the manuscript. MS performed the literature search, screening, data extraction and data analysis, review of articles, corrected the manuscript, IS reviewed the study results and final draft, KN reviewed the introduction and final draft. RR reviewed and corrected the manuscript. AL performed the literature search, screening, data extraction and data analysis. AH corrected the final manuscript. BM reviewed and corrected the final draft.

