## Supplemental Table 1 for "Implementation of Point of Care HIV Viral load monitoring for people living with HIV in Low and Middle-income Countries: A systematic review on implementation research outcomes"

S1 Table Search Strategy

Keywords 3/06/2024

1. Point of care test
2. HIV
3. Viral load
4. Acceptability
5. Feasibility
6. Appropriateness
7. Cost
8. Adoption
9. Fidelity
10. Treatment monitoring

PubMed search strategy (55hits)

| | |
|---|---|
| 1. | "Point of care testing" [mesh] OR "Testing, Point-of-Care*"[tiab] OR "Point-Of-Care Diagnostic Testing*" [tiab] OR "Point Of Care Diagnostic Testing*"[tiab] OR "Point-Of-Care Diagnostic Tests"[tiab] OR "Point Of Care Diagnostic Tests*"[tiab] OR "Point-Of-Care Diagnostic Test*"[tiab] OR "Point Of Care Tests*"[tiab] OR "Point-Of-Care Test*"[tiab] OR "Point Of Care Test*"[tiab] OR "Test, Point-Of-Care*"[tiab] OR "Tests, Point-Of-Care*"[tiab] OR "Point of Care Testing"[tiab] "Point-Of-Care Diagnostics*"[tiab] OR "Point Of Care Diagnostics*"[tiab] OR "Point-Of-Care Diagnostic*"[tiab] OR "Bedside Testing*"[tiab] OR "Testing, Bedside*" [tiab] OR POC [tiab] |
| 2. | "HIV "[mesh] OR "HIV infections"[mesh] OR "Human Immunodeficiency Virus"[tiab] OR "Immunodeficiency Virus, Human*"[tiab] OR "Immunodeficiency Viruses, Human*"[tiab] OR "Virus, Human Immunodeficiency*"[tiab] OR "Viruses, Human Immunodeficiency*"[tiab] OR "Human Immunodeficiency Viruses"[tiab] OR "Human T Cell Lymphotropic Virus Type III*"[tiab] OR "Human T-Cell Lymphotropic Virus Type III*"[tiab] OR "Human T-Cell Leukemia Virus Type III*"[tiab] OR "Human T Cell Leukemia Virus Type III*"[tiab] OR "LAV-HTLV-III"[tiab] OR "Virus, Lymphadenopathy Associated*"[tiab] OR "Viruses, Lymphadenopathy Associated*"[tiab] OR "Human T Lymphotropic Virus Type III*"[tiab] OR "Human T-Lymphotropic Virus Type III*"[tiab] OR "Virus, AIDS*"[tiab] OR "Viruses, AIDS*"[tiab] OR "Acquired Immune Deficiency Syndrome Virus*"[tiab] OR "Acquired Immunodeficiency Syndrome Virus"[tiab] OR "HTLV-III"[tiab] OR "HIV Infection"[tiab] OR "HIV coinfection"[tiab] OR HIV[tiab] |
| 3. | "Viral load" [mesh] OR "Load, Viral" [tiab] OR "Viral Burden*" [tiab] OR "Burden, Viral*" [tiab] OR "Virus Titer*" [tiab] OR "Titer, Virus*" [tiab] |
| 4. | "Patient Acceptance of Health Care" [mesh] OR "Health Care Utilization"[tiab] OR "Utilization, Health Care"[tiab] OR "Patient Acceptance of Healthcare"[tiab] OR "Health Care Seeking Behavior"[tiab] OR "Acceptability of Health Care"[tiab] OR "Health Care Acceptability"[tiab] OR "Acceptability of Healthcare"[tiab] OR Patient Acceptance of Health Care[tiab] OR Acceptability[tiab] |

| | |
|---|---|
| 5. | "Feasibility studies" [mesh] OR "Feasibility Study" [tiab] OR "Studies, Feasibility" [tiab] OR "Study, Feasibility" [tiab] OR feasibility[tiab] |
| 6. | "Continuity of Patient Care"[mesh] OR "program evaluation"[mesh] OR "Care Continuity, Patient"[tiab] OR "Patient Care Continuity"[tiab] OR "Continuum of Care"[tiab] OR "Care Continuum"[tiab] OR "Continuity of Care"[tiab] OR "Care Continuity"[tiab] OR "Evaluation, Program"[tiab] OR "Evaluations, Program"[tiab] OR "Program Evaluations"[tiab] OR "Program Sustainabilit*"[tiab] OR "Sustainability Program"[tiab] OR "Program Effectiveness"[tiab] OR "Effectiveness, Program"[tiab] OR "Program Appropriateness"[tiab] OR "Appropriateness, Program"[tiab] OR "Penetration"[tiab] |
| 7. | "Cost-Benefit Analysis" [mesh] OR "Analyses, Cost-Benefit"[tiab] OR "Analysis, Cost-Benefit"[tiab] OR "Cost-Benefit Analyses" [tiab] OR "Cost Benefit Analysis"[tiab] OR "Analyses, Cost Benefit"[tiab] OR "Analysis, Cost Benefit" [tiab] OR "Cost Benefit Analyses" [tiab] OR "Cost Effectiveness"[tiab] OR "Effectiveness, Cost"[tiab] OR "Cost-Benefit Data" [tiab] OR "Cost Benefit Data"[tiab] OR "Data, Cost-Benefit"[tiab] OR "Cost-Utility Analysis"[tiab] OR "Analyses, Cost-Utility" [tiab] OR "Analysis, Cost-Utility"[tiab] OR "Cost Utility Analysis"[tiab] OR "Cost-Utility Analyses"[tiab] OR "Economic Evaluation"[tiab] OR "Economic Evaluations"[tiab] OR "Evaluation, Economic"[tiab] OR "Evaluations, Economic"[tiab] OR "Marginal Analysis"[tiab] OR "Analyses, Marginal" [tiab] OR "Analysis, Marginal" [tiab] OR "Marginal Analyses" [tiab] OR "Cost Benefit"[tiab] OR "Costs and Benefits"[tiab] OR "Benefits and Costs"[tiab] OR "Cost-Effectiveness Analysis"[tiab] OR "Analysis, Cost-Effectiveness"[tiab] OR "Cost Effectiveness Analysis"[tiab] OR "Cost"[tiab] |
| 8. | "Facilities and services Utilization" [mesh] OR "Facilities Utilization" [tiab] OR "Services Utilization" [tiab] OR "Services Utilizations" [tiab] OR "Adoption" |
| 9. | fidelity |
| 10. | treatment monitoring |
| 11. | #1 AND #2 AND #3 AND (#4 OR #5 OR #6 OR #7 OR #8 OR #9 OR #10) |

Scopus search strategy (242 hits)

| | |
|---|---|
| 1. | TITLE-ABS-KEY ( point AND of AND care AND testing ) |
| 2. | TITLE-ABS-KEY ( hiv ) |
| 3. | TITLE-ABS-KEY ( viral AND load ) |
| 4. | TITLE-ABS-KEY ( acceptability ) |
| 5. | TITLE-ABS-KEY ( adoption ) |
| 6. | TITLE-ABS-KEY ( appropriateness ) |
| 7. | TITLE-ABS-KEY ( cost ) |
| 8. | TITLE-ABS-KEY ( feasibility ) |
| 9. | TITLE-ABS-KEY ( fidelity ) |

| | |
|---|---|
| 10. | TITLE-ABS-KEY ( sustainability ) |
| 11. | TITLE-ABS-KEY ( treatment AND monitoring ) |
| 12. | 1 AND 2 AND 3<br><br>( TITLE-ABS-KEY ( point AND of AND care AND testing ) ) AND ( TITLE-ABS-KEY ( hiv ) ) AND ( TITLE-ABS-KEY ( viral AND load ) ) |
| 13. | 4 OR 5 OR 6 OR 7 OR 8 OR 9 OR 10 OR 11<br><br>( TITLE-ABS-KEY ( acceptability ) ) OR ( TITLE-ABS-KEY ( adoption ) ) OR ( TITLE-ABS-KEY ( appropriateness ) ) OR ( TITLE-ABS-KEY ( cost ) ) OR ( TITLE-ABS-KEY ( feasibility ) ) OR ( TITLE-ABS-KEY ( fidelity ) ) OR ( TITLE-ABS-KEY ( sustainability ) ) OR ( TITLE-ABS-KEY ( treatment AND monitoring ) ) |
| 14. | 12 AND 13<br><br>( ( TITLE-ABS-KEY ( point AND of AND care AND testing ) ) AND ( TITLE-ABS-KEY ( hiv ) ) AND ( TITLE-ABS-KEY ( viral AND load ) ) ) AND ( ( TITLE-ABS-KEY ( acceptability ) ) OR ( TITLE-ABS-KEY ( adoption ) ) OR ( TITLE-ABS-KEY ( appropriateness ) ) OR ( TITLE-ABS-KEY ( cost ) ) OR ( TITLE-ABS-KEY ( feasibility ) ) OR ( TITLE-ABS-KEY ( fidelity ) ) OR ( TITLE-ABS-KEY ( sustainability ) ) OR ( TITLE-ABS-KEY ( treatment AND monitoring ) ) ) |

Cochrane search strategy (16 hit)

| | |
|---|---|
| 1. | ("Point of care test"):ti,ab,kw |
| 2. | ("HIV"):ti,ab,kw |
| 3. | ("viral load"):ti,ab,kw |
| 4. | #1 AND #2 AND #3 |
| 5. | ("Acceptability"):ti,ab,kw |
| 6. | ("Adoption"):ti,ab,kw |
| 7. | ("Appropriateness"):ti,ab,kw |
| 8. | ("Cost"):ti,ab,kw |
| 9. | ("Feasibility"):ti,ab,kw |
| 10. | ("Fidelity"):ti,ab,kw |
| 11. | ("Sustainability"):ti,ab,kw |
| 12. | ("Treatment monitoring"):ti,ab,kw |
| 13. | #5 OR #6 OR #7 OR #8 OR #9 OR #10 OR #11 OR #12 |
| 14. | #4 AND #13 |
