## Supplemental Table 2 for "Implementation of Point of Care HIV Viral load monitoring for people living with HIV in Low and Middle-income Countries: A systematic review on implementation research outcomes"

### PRISMA 2020 Checklist

| Section and Topic | Item # | Checklist item | Reported on page # |
|---|---|---|---|
| **TITLE** | | | |
| Title | 1 | Identify the report as a systematic review. | 1 |
| **ABSTRACT** | | | |
| Abstract | 2 | Provide a structured summary including, as applicable: background; objectives; data sources; study eligibility criteria, participants, and interventions; study appraisal and synthesis methods; results; limitations; conclusions and implications of key findings; systematic review registration number. | 1-2 |
| **INTRODUCTION** | | | |
| Rationale | 3 | Describe the rationale for the review in the context of existing knowledge. | 3-4 |
| Objectives | 4 | Provide an explicit statement of the objective(s) or question(s) the review addresses. | 5 |
| **METHODS** | | | |
| Eligibility criteria | 5 | Specify the inclusion and exclusion criteria for the review and how studies were grouped for the syntheses. | 7-8 |
| Information sources | 6 | Specify all databases, registers, websites, organisations, reference lists and other sources searched or consulted to identify studies. Specify the date when each source was last searched or consulted. | 6 |
| Search strategy | 7 | Present the full search strategies for all databases, registers and websites, including any filters and limits used. | 6, S1 |
| Selection process | 8 | Specify the methods used to decide whether a study met the inclusion criteria of the review, including how many reviewers screened each record and each report retrieved, whether they worked independently, and if applicable, details of automation tools used in the process. | 6-7 |
| Data collection process | 9 | Specify the methods used to collect data from reports, including how many reviewers collected data from each report, whether they worked independently, any processes for obtaining or confirming data from study investigators, and if applicable, details of automation tools used in the process. | 6-7 |
| Data items | 10 | List and define all outcomes for which data were sought. Specify whether all results that were compatible with each outcome domain in each study were sought (e.g. for all measures, time points, analyses), and if not, the methods used to decide which results to collect. | 7 |
| Study risk of bias assessment | 11 | Specify the methods used to assess risk of bias in the included studies, including details of the tool(s) used, how many reviewers assessed each study and whether they worked independently, and if applicable, details of automation tools used in the process. | 6-7 |
| **RESULTS** | | | |
| Study selection | 12 | Describe the results of the search and selection process, from the number of records identified in the search to the number of studies included in the review, ideally using a flow diagram. | 9-10 |
| | | Cite studies that might appear to meet the inclusion criteria, but which were excluded, and explain why they were excluded. | 9-10 |
| Study characteristics | 13 | Cite each included study and present its characteristics. | 11 |
| Risk of bias in studies | 14 | Present assessments of risk of bias for each included study. | 6-7 |
| **DISCUSSION** | | | |
| Discussion | 15 | Provide a general interpretation of the results in the context of other evidence. | 25-28 |
| | | Discuss any limitations of the evidence included in the review | 26 |

### PRISMA 2020 Checklist

| Section and Topic | Item # | Checklist item | Reported on page # |
|---|---|---|---|
| | | Discuss any limitations of the review processes used. | 26 |
| | 16 | Discuss implications of the results for practice, policy, and future research. | 27 |
| **OTHER INFORMATION** | | | |
| Registration | 17 | Provide registration information for the review, including register name and registration number, or state that the review was not registered. | 2 |
| Support | 18 | Describe sources of financial or non-financial support for the review, and the role of the funders or sponsors in the review. | 28 |
| Competing interests | 19 | Declare any competing interests of review authors. | 28 |
| Availability of data, code and other materials | 20 | Report which of the following are publicly available and where they can be found: template data collection forms; data extracted from included studies; data used for all analyses; analytic code; any other materials used in the review. | 28 |

*From:* Page MJ, McKenzie JE, Bossuyt PM, Boutron I, Hoffmann TC, Mulrow CD, et al. The PRISMA 2020 statement: an updated guideline for reporting systematic reviews. BMJ 2021;372:n71. doi: 10.1136/bmj.n71

For more information, visit: http://www.prisma-statement.org/
