## Supplemental Table 3 for "Implementation of Point of Care HIV Viral load monitoring for people living with HIV in Low and Middle-income Countries: A systematic review on implementation research outcomes"

### S3 Table Reasons for exclusion.

A total of 9 studies were excluded. Reasons for exclusion

| First Author | Year of publication | Title | Reason for exclusion |
|---|---|---|---|
| Ritchie | 2014 | SAMBA HIV semiquantitative test, a new point of care viral load monitoring assay for resource-limited settings | Wrong outcomes |
| Moirana | 2022 | Evaluation of HIV viral load turnaround time in Moshi, Tanzania | Wrong intervention |
| Nakyanzi | 2024 | It soothes your heart. A Multimethod Study Exploring Acceptability of Point of Care Viral Load Testing among Ugandan Pregnant and Postpartum Women Living with HIV | Wrong study design |
| Dorward | 2018 | Point of care viral load testing and differentiated HIV care | Wrong intervention |
| Stevens | 2014 | Feasibility of HIV point of care tests for resource-limited settings: Challenges and solutions | Wrong study design |
| Moyo | 2016 | Point of care Cepheid Xpert HIV-1 Viral Load Test in Rural African Communities Is Feasible and Reliable | Wrong outcomes |
| Qian | 2022 | After viral load testing, I get my results, so I get to know which path my life is taking me. qualitative insights on routine centralised and point-of-care viral load testing in western Kenya from the Opt4Kids and Opt4Mamas studies | Wrong outcome |
| Avram | 2019 | Point of care HIV viral load in pregnant women without prenatal care: a cost-effectiveness analysis | Wrong study design |
| Drain | 2019 | Point of Care HIV Viral load Testing: an Essential Tool for a Sustainable Global HIV/AIDS Response | Wrong intervention |
