## Supplemental Table 4 for "Implementation of Point of Care HIV Viral load monitoring for people living with HIV in Low and Middle-income Countries: A systematic review on implementation research outcomes"

S4 Table Newcastle Ottawa scale. The quality assessment of the non-randomized studies

| Study Author (year) | Outcome | A. Selection (maximum of four stars) | | | | Comparability (maximum of two stars) | C. Outcome (maximum of three stars) | | | Total (maximum of nine stars) |
|---|---|---|---|---|---|---|---|---|---|---|
| | | 1. Representativeness of the exposed cohort | 2. Selection of the non-exposed cohort | 3. Ascertainment of exposure | 4. Demonstration that outcome of interest was not present at start of study | 1. Comparability of cohort based on the design or analysis | 1. Assessment of outcome | 2. Was follow-up long enough for outcomes to occur | 3. Adequacy of follow-up of cohorts | |
| Boyce (2023) | Feasibility | ★ | ☆ | ★ | ★ | ☆ | ★ | ★ | ★ | 6 |
| Tembo (2022) | Acceptability | ★ | ☆ | ★ | ★ | ☆ | ★ | ★ | ★ | 6 |
| Ganesh (2021) | Feasibility | ★ | ☆ | ★ | ☆ | ★ | ★ | ★ | ★ | 6 |
| Wang (2021) | Feasibility, fidelity | ★ | ☆ | ★ | ☆ | ★ | ★ | ★ | ★ | 6 |
| Broucker (2021) | Cost | ★ | ☆ | ★ | ★ | ★ | ★ | ★ | ☆ | 6 |
| Boeke (2021) | Appropriateness, Sustainability | ★ | ★ | ★ | ★ | ★ | ★ | ★ | ★ | 8 |
| Bulterys (2021) | Cost, feasibility | ★ | ★ | ★ | ★ | ★ | ★ | ★ | ★ | 8 |
| Gueguen (2021) | Adoption, Feasibility, penetration, sustainability | ★ | ★ | ★ | ★ | ★ | ★ | ★ | ☆ | 7 |
| Sharma (2021) | Acceptability, sustainability | ★ | ★ | ★ | ★ | ★ | ★ | ★ | ★ | 8 |
| Kufa (2020) | Feasibility, penetration | ★ | ☆ | ★ | ★ | ★ | ★ | ★ | ☆ | 6 |
| Villa (2020) | Acceptability | ★ | ★ | ★ | ★ | ★ | ★ | ★ | ★ | 8 |
| Vasconcellos (2020) | Feasibility | ★ | ☆ | ★ | ★ | ★ | ★ | ★ | ★ | 7 |

S4 Table Newcastle Ottawa scale. The quality assessment of the non-randomized studies

| Study Author (year) | Outcome | A. Selection (maximum of four stars) | | | | Comparability (maximum of two stars) | C. Outcome (maximum of three stars) | | | Total (6maximum of nine stars) |
|---|---|---|---|---|---|---|---|---|---|---|
| | | 1. Representativeness of the exposed cohort | 2. Selection of the non-exposed cohort | 3. Ascertainment of exposure | 4. Demonstration that outcome of interest was not present at start of study | 1. Comparability of cohort based on the design or analysis | 1. Assessment of outcome | 2. Was follow-up long enough for outcomes to occur | 3. Adequacy of follow-up of cohorts | |
| Msimango (2020) | Acceptability, Appropriateness | ★ | ★ | ★ | ☆ | ★ | ★ | ☆ | ★ | 6 |
| Girdwood (2020) | Cost, Sustainability | ★ | ★ | ★ | ☆ | ★ | ★ | ★ | ★ | 7 |
| Girdwood (2019) | Adoption, Appropriateness | ★ | ★ | ★ | ☆ | ★ | ★ | ★ | ★ | 7 |
| Nicholas (2019) | Acceptability | ★ | ★ | ★ | ☆ | ★ | ★ | ☆ | ★ | 6 |
| Simeon (2019) | Cost | ★ | ★ | ★ | ★ | ★ | ★ | ★ | ★ | 8 |
| Necker (2019) | Cost | ★ | ★ | ★ | ★ | ★ | ★ | ★ | ★ | 8 |
| Ndlovu (2018) | Adoption, Appropriateness, cost | ☆ | ★ | ☆ | ☆ | ★ | ★ | ☆ | ☆ | 3 |
| Engel (2015) | Acceptability, Adoption, cost, Feasibility, penetration | ★ | ★ | ★ | ★ | ★ | ★ | ☆ | ★ | 7 |
| Engel (2017) | Acceptability, Fidelity, sustainability | ★ | ★ | ★ | ★ | ★ | ★ | ☆ | ★ | 7 |
| Rasti (2017) | Acceptability, Adoption | ★ | ★ | ★ | ☆ | ★ | ★ | ☆ | ★ | 6 |
| Estill (2013) | Cost, Sustainability | ☆ | ☆ | ★ | ☆ | ★ | ★ | ★ | ☆ | 4 |

S4 Table Newcastle Ottawa scale. The quality assessment of the non-randomized studies

| Study Author (year) | Outcome | A. Selection (maximum of four stars) | | | | Comparability (maximum of two stars) | C. Outcome (maximum of three stars) | | | Total (6maximum of nine stars) |
|---|---|---|---|---|---|---|---|---|---|---|
| | | 1. Representativeness of the exposed cohort | 2. Selection of the non-exposed cohort | 3. Ascertainment of exposure | 4. Demonstration that outcome of interest was not present at start of study | 1. Comparability of cohort based on the design or analysis | 1. Assessment of outcome | 2. Was follow-up long enough for outcomes to occur | 3. Adequacy of follow-up of cohorts | |
| Reif (2022) | Acceptability | ★ | ★ | ★ | ★ | ★★ | ★ | ★ | ★ | 9 |
| Drain (2020) | Acceptability, Feasibility | ★ | ★ | ★ | ★ | ★★ | ★ | ★ | ★ | 9 |
